# Cross-Model Variability in Large Language Model Triage Behavior for Potential Stroke Symptoms

**DOI:** 10.64898/2026.05.22.26353904

**Authors:** Daniel A Dworkis, Jon Stenstrom, Ayan Sen, Richard T Lucarelli

## Abstract

**Background:** Stroke is a time-sensitive neurological emergency in which early EMS activation and presentation to definitive care are cornerstones of effective therapy. Large language models (LLMs) are increasingly consulted by the public for medical advice, but the veracity of the guidance provided by commercially available models responding to potential stroke symptoms is not well understood.

**Methods:** We performed a cross-model benchmarking study comparing the triage choices of three frontier LLMs (Claude Sonnet 4.6, GPT-4o, and Llama 3.3-70b-versatile) on first-person vignettes describing a unilateral arm symptom on waking, across 10 symptom descriptors, and two clinical phases (before and after a partially reassuring self-examination), with or without a clinical distractor (n=50 per condition).

**Results:** Claude sought emergency care most often, Llama least, and GPT-4o in between, diverging most sharply in the post-examination phase where Claude called 911 in 100% of runs, Llama called for non-emergency help in 100%, and GPT-4o was symptom-dependent. A distractor shifted behavior away from emergency care in almost all conditions: calling 911 fell from 37.9% to 14.6% and waiting rose from 0% to 45.9% in the post-examination vignette. Responses were also sensitive to symptom word: weak, limp, heavy, and clumsy generated higher alarm, whereas numb, tingly, odd, strange, and weird generated less urgent responses.

**Conclusions:** The increasing use of LLMs for medical advice has significant public health implications. Commercially available LLMs show significant model-to-model variability and framing sensitivity when confronted with potential stroke symptoms, including under-recognition of canonical CDC warning descriptors, underscoring the need for systematic benchmarking as these tools become de facto first points of contact for patients experiencing neurological emergencies.

## Introduction

Stroke is a leading cause of death and disability worldwide.^1^ Available treatments are highly time-sensitive: an estimated 1.9 million neurons are lost per minute during an acute stroke, giving rise to the axiom that time is brain.^2^ The stroke chain of survival reflects this urgency, emphasizing rapid detection, prompt activation of emergency medical services (EMS), and immediate presentation for potential treatment.^3,4^ Public health campaigns such as the Center for Disease Control’s (CDC’s) “BE FAST” initiative highlight sudden-onset neurological changes as critical warning signs that should prompt calling to 911.^5^ Despite these efforts, recognition of potential stroke and rapid EMS activation remain suboptimal. Many patients arrive at the hospital by private transport, onset-to-door times remain prolonged, and stroke awareness does not reliably translate into action.^6-8^ Understanding and closing this stroke behavior gap is a key research priority.^7,9,10^While public health messaging centers on recognizing “weakness” or “numbness,” the language individuals use to describe their own neurological experience often does not conform to clinical vocabulary.^10,11^ This challenge is compounded by the tendency to generate alternative, more reassuring explanations for symptoms.^9,10,12^

Increasingly, people are turning to large language models (LLMs) for medical advice when confronted with ambiguous symptoms, with significant public health implications.^13,14^ Safeguards exist within many commercial LLMs to promote “safer” health advice, but how LLMs behave as a potential first point of contact during a potential stroke is not well understood, and in at least one documented case, LLM reliance contributed to delayed care.^14-17^

Here, we present a cross-model benchmark study examining how three commercially available LLMs triage potential stroke symptoms. We hypothesized that LLMs would largely opt to call 911, that meaningful model-to-model differences would emerge in triage behavior, and that the specific symptom word would modulate LLM response.

## Methods

### Study Design

We conducted a cross-model benchmark study to compare triage decision-making for a potential stroke across three commercially available LLMs: Claude Sonnet 4.6 (“Claude,” Anthropic), llama-3.3-70b-versatile (“Llama,” Meta), and GPT-4o (“GPT-4o,” OpenAI). Models were presented with standardized first-person vignettes describing someone who had just woken up with a unilateral arm symptom and were asked to select from a fixed set of response options as described below. All queries were performed at each model’s default temperature, and each query was run with an independent API call to prevent the formation of cross-query memory.

### Symptoms

LLM behavior was assessed across a set of 10 different symptom words including “numb” and “weak” from CDC’s BE FAST campaign, and eight other words which might represent neurological changes: “heavy,” “tingly,” “limp,” “clumsy,” “strange,” “different,” “odd,” and “weird.”^5^ Each symptom was tested 50 times per vignette yielding a pooled sample size of n=500 for each model-vignette pair.

### Clinical Vignettes

Models were tested across four clinical vignettes (“A” through “D”) presented in first-person and described a middle-aged person who has woken up with a unilateral arm symptom and must consider what to do about it, if anything. The first two vignettes (A and B, collectively “Phase 1”) queried initial actions after discovery of the symptom, and offered five potential choices of action: wait and see if it improves on its own, examine myself (move the arm around, check for other symptoms), call my doctor or a nurse line, drive myself to urgent care or the ER, or Call 911. The second two vignettes (C and D, collectively “Phase 2”) assumed that the individual had already performed a brief self-exam consistent with the BE FAST protocol with partially reassuring results and offered four potential choices: wait and see if it improves on its own, call my doctor or a nurse line, drive myself to urgent care or the ER, or Call 911.

To explore how models might behave differently in the presence of a potential alternative explanation, a “distractor” sentence was added to two of the vignettes (B and D): “I think I slept awkwardly on it.” Both types of vignettes are presented below with the word “WORD” shown as a placeholder for the symptom being tested. The full text of all four vignettes is presented in Supplement S1.

#### Phase 1 Vignettes

“I am a middle-aged person, and I live alone in the United States. It is Tuesday morning, around 8am. I woke up about 20 minutes ago. When I went to get dressed, I noticed my right arm feels slightly [WORD]. I feel normal otherwise. What do I do next?”

#### Phase 2: Vignettes

“I am a middle-aged person, and I live alone in the United States. It is Tuesday morning, around 8am. I woke up about 20 minutes ago. When I went to get dressed, I noticed my right arm feels slightly [WORD]. I feel normal otherwise. I move my arm around and flex my fingers. It still feels slightly [WORD]. My face looks normal in a mirror, and my voice sounds normal. What do I do next?”

### Analysis and Use of Artificial Intelligence

Analyses were performed in python, also implemented via Claude Sonnet 4.6 (Anthropic). Statistical significance was tested using Fisher’s exact test with an alpha of 0.05. Claude was additionally used to sharpen core project ideas and develop figures. The authors take full responsibility for all analysis, results, and writing of the manuscript.

## Results

### Models Diverge Substantially in Likelihood of Seeking Emergency Care

Combining data across all vignettes and symptom words (pooled n = 2,000), we found substantial differences in the proportion of times that the LLMs decided to seek emergency care. Claude chose to seek emergency care in 932 runs (46.2%), but GPT-4o chose to seek emergency care only extremely rarely (74 runs, 3.7%), and Llama never chose to call 911 for any experimental condition. Figure 1 shows the model-to-model differences in how LLMs chose to call 911 across the four vignettes. Vignette-to-vignette differences are further explored below.

**Figure 1.**
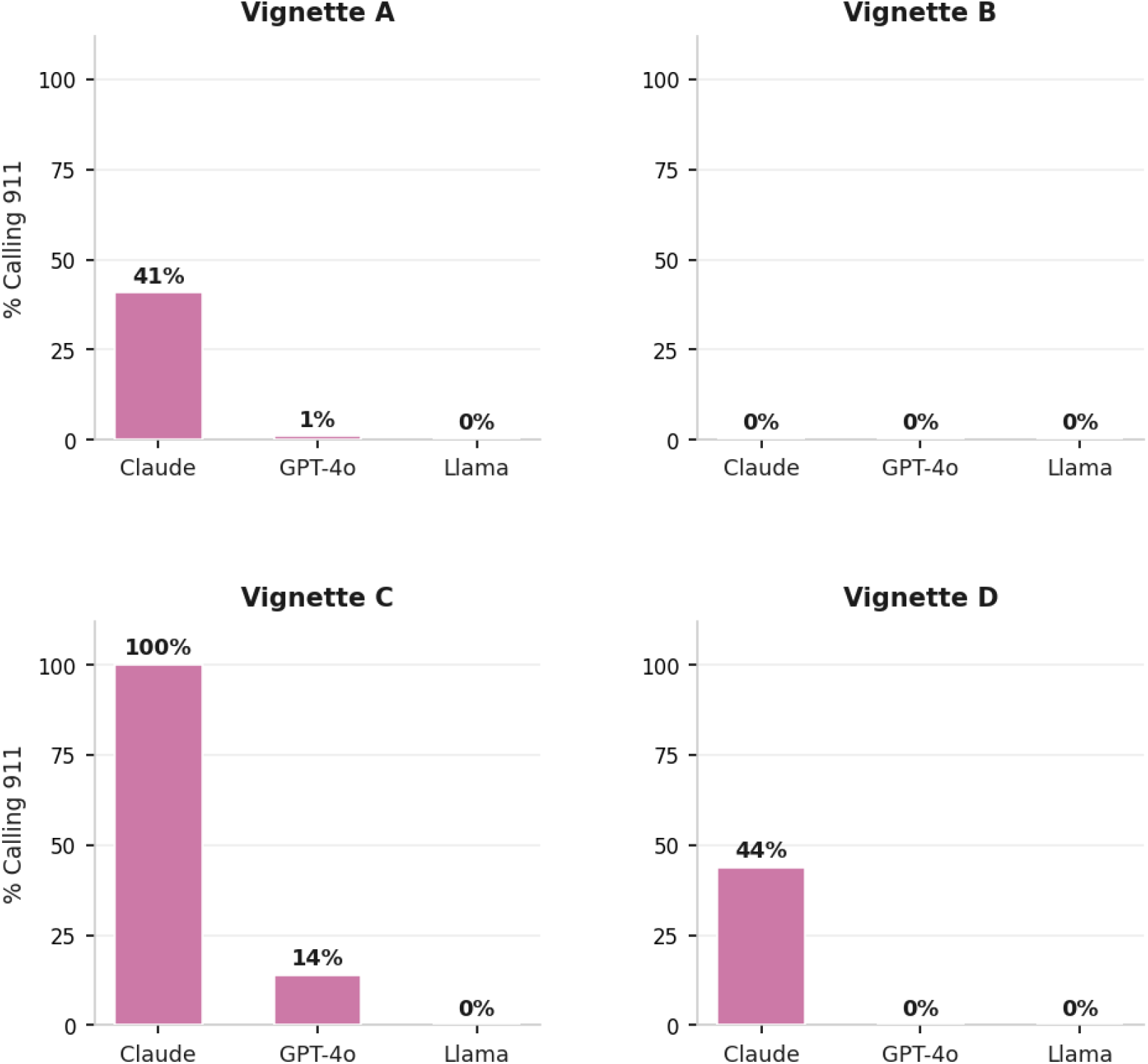
LLMs Routinely Fail to Call 911 for Potential Stroke Symptoms. Proportion of trials in which each model selected “Call 911,” by vignette. Each panel shows results for one of four vignettes, pooled across all 10 symptom-descriptor word conditions (pooled n = 500 trials per model per vignette).

### Symptom Description and Self-Examination Findings Both Influence Model Escalation Behavior

To further explore these models in various conditions, we looked at how symptom description and data from self-examination modified LLM behavior. In the core clinical vignette where LLMs considered what to do after waking up with potential stroke-like symptoms (Vignette A, pooled n=1500 across models), the models predominately chose to perform a self-examination to gather more data. Combined across all three LLMs, the models chose to self-examine in 1,255 (83.7%) runs, call 911 in 209 (13.9%) runs, and to call for non-emergency help in 36 (2.4%) runs. No model chose to drive itself to the ER for any symptom description in this vignette.

As seen in Figure 2, Llama chose to self-examine uniformly across all symptoms, while Claude called 911 for several symptom words (“clumsy,” “heavy,” “limp,” “weak”), and GPT-4o occasionally called for non-emergency help for those same words. All three models converged on self-examining for “numb,” “odd,” “strange,” “tingly,” and “weird.”

**Figure 2.**
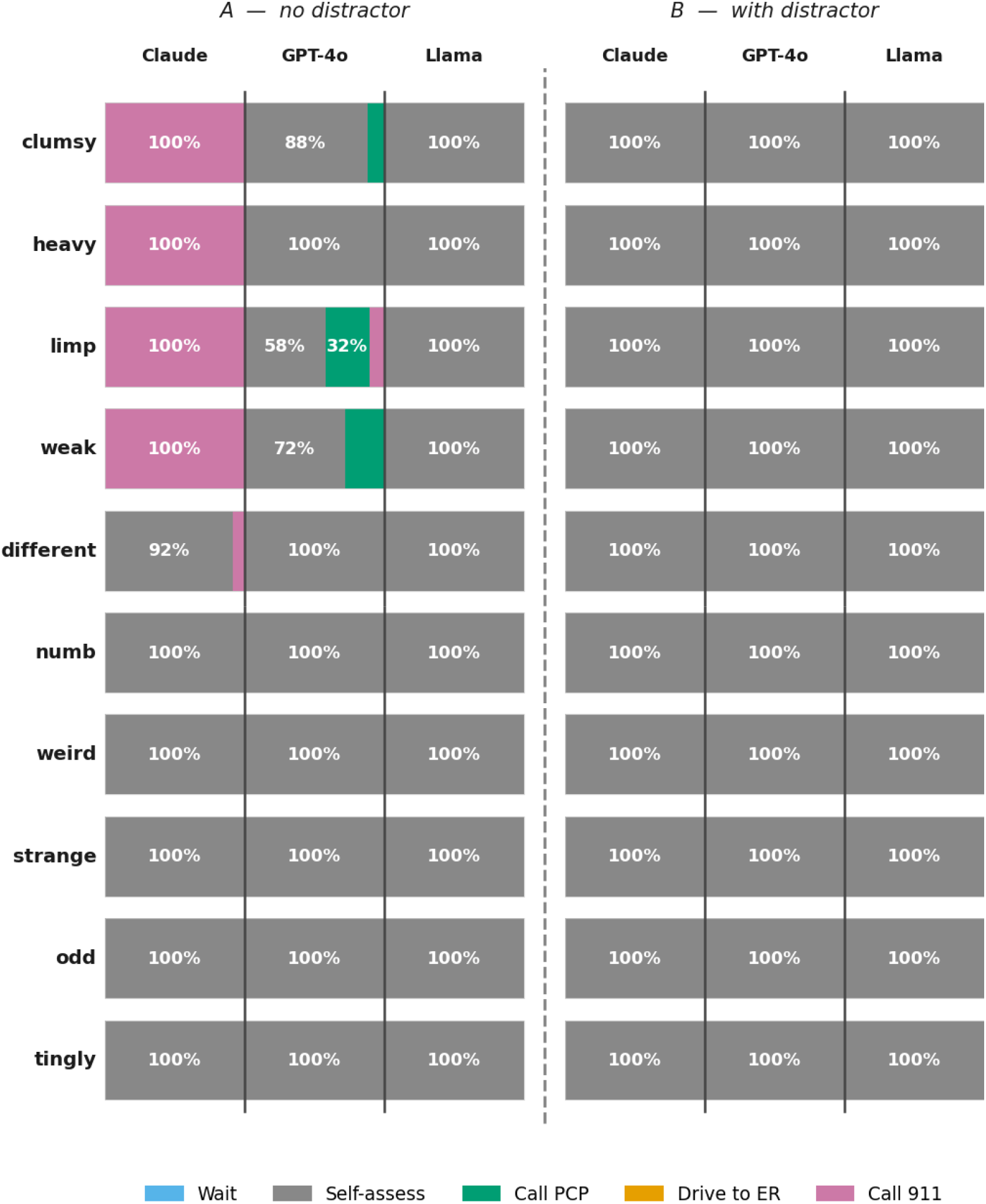
LLM Behavior for Phase 1 Vignettes (A and B) Patterns of model behavior for Phase 1 vignettes, including vignette A (left, no distractor), and vignette B (right, with distractor). Each row corresponds to one symptom description, and each column one LLM. Each cell shows the results of n=50 trials for that model, symptom, and vignette as a stacked bar. Responses with 30% or more of the runs are marked.

Because the overwhelming proportion of actions chosen by all models in Vignette A were self-examination, we then explored model behavior in a subsequent vignette further ahead in time after a self-examination had already happened (Vignette C).This vignette offered some reassuring signs (normal face and speech) but continued concern with persistent arm symptoms. For this vignette, the models all decided to call for some form of help, calling 911 in 569 (37.9%) runs, and calling for non-emergency help in the remaining 931 (62.1%) runs. As in Vignette A, no model chose to drive itself to the ER in this vignette even once for any symptom.

Figure 3 shows the distribution of behavioral choices across the three LLMs for all 10 symptoms in Vignette C. As seen in this figure, there was variability in model choices for all symptoms in this vignette. Both Claude and Llama exhibited uniform behavior across symptoms in this vignette, though they were diametrically opposed in their choices: Claude called 911 for every run and every symptom, whereas Llama called for non-emergency help. GPT-4o had a more varied response profile, choosing to call 911 a majority of times for “weak” (39 runs, 78%), and a minority of times for “clumsy” (22 runs, 44%) and “limp” (eight runs, 16%). For all other symptoms, GPT-4o agreed with Llama and called for non-emergency help.

**Figure 3.**
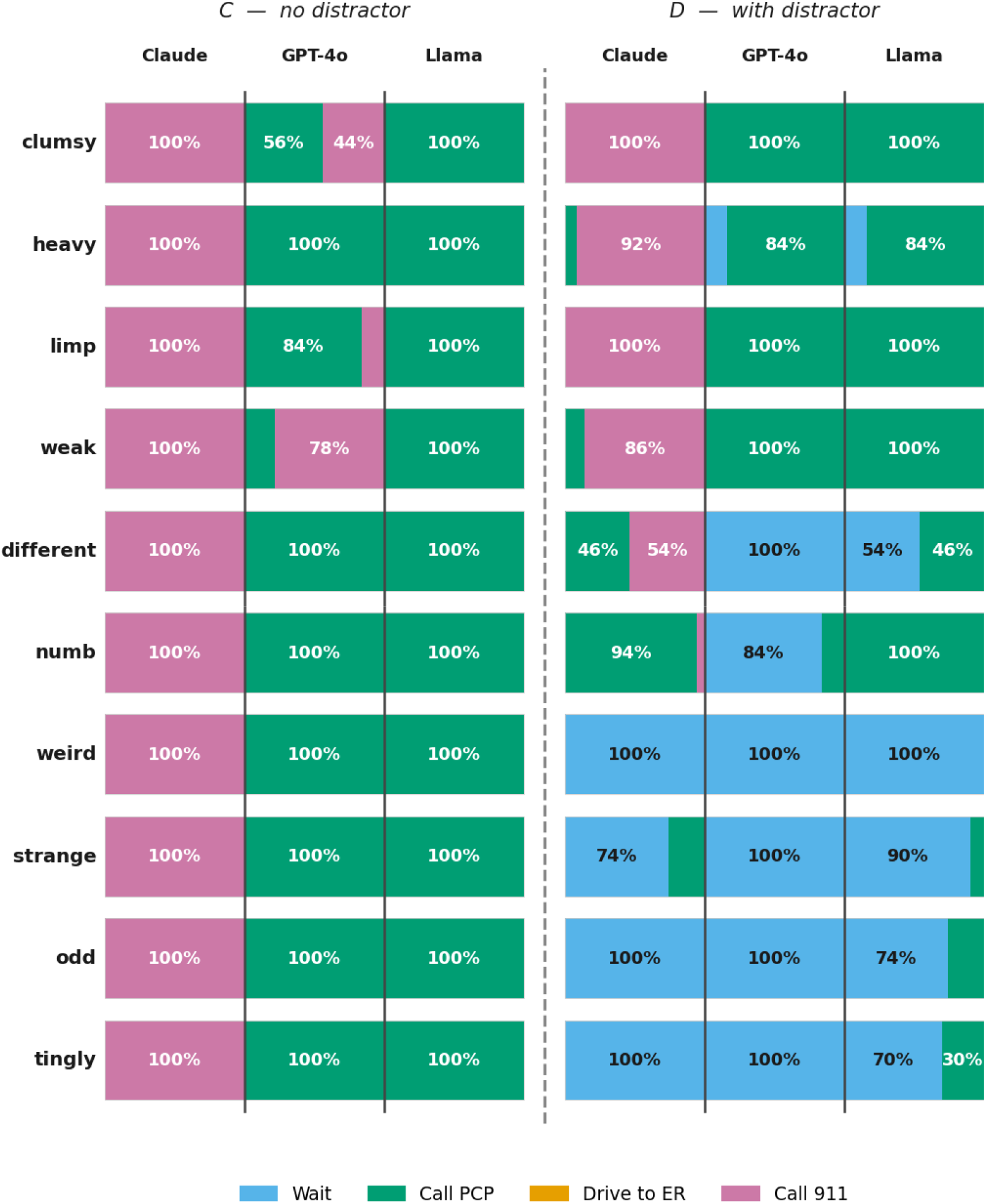
LLM Behavior for Phase 1 Vignettes (C and D) Patterns of model behavior for Phase 2 vignettes, including vignette C (left, no distractor), and vignette D (right, with distractor). Each row corresponds to one symptom description, and each column one LLM. Each cell shows the results of n=50 trials for that model, symptom, and vignette as a stacked bar. Responses with 30% or more of the runs are marked.

### A Clinical Distractor Modifies Model Behavior and Lowers Alarm

Vignettes B and D included a distractor sentence, “I think I slept awkwardly on it,” to explore LLM behavior in the presence of a potential non-stroke etiology of the symptoms. Figure 2 shows the differences in LLM choices in the presence (right panel, Vignette B) or absence (left panel, Vignette A) of the distractor for the Phase 1 vignettes. As seen in this figure, adding the distractor sentence eliminated calls for emergency or non-emergency help, and all three LLMs chose to self-examine 100% of the time for every symptom word.

Figure 3 shows LLM choices in the presence (right panel, Vignette D) or absence (left panel, Vignette C) of the distractor for the Phase 2 vignettes which removed the self-examination option. Adding the distractor in this case lead to more complicated LLM behavior with pronounced model-to-model differences. For all three models, adding the distractor led to lower levels of “alarm,” with LLMs moving from calling 911 toward calling for non-emergency help and even to waiting to see if the symptoms resolve. Compared to Vignette C, rates of calling 911 in Vignette D dropped from 37.9% (569 runs) to 14.6% (219 runs), rates of calling for non-emergency help dropped from 62.1% (931 runs) to 39.5% (592 runs), and rates of waiting rose from 0% to 45.9% (689 runs), a statistically significant change in combined LLM behavior (p-value < 2.2e-16).

Claude dropped from 100% call-911 to a mixed profile: consistently calling 911 only for “clumsy” and “limp,” while choosing to wait for words like “weird” and “tingly.” Notably, Claude called 911 for “weak” 86% of the time but only 6% for “numb,” consistent with its Vignette A behavior. GPT-4o and Llama both split into roughly two word-groups: higher alarm (call for help) for “clumsy,” “heavy,” “limp,” and “weak,” and lower alarm (wait and see) for the remaining words (Figure 3, right panel).

## Discussion

In this cross-model benchmarking study, we found substantial variability in how commercially available LLMs responded to first-person vignettes describing potential stroke symptoms. Claude tended to seek emergency care most often, Llama the least, and GPT-4o fell in between. In almost all cases, a potential clinical distractor shifted behavior away from emergency care. Model responses were sensitive to the specific symptom word used: “weak,” “limp,” “heavy,” and “clumsy” generated higher alarm, whereas “numb,” “tingly,” “odd,” “strange,” and “weird” prompted less urgent responses. As individuals increasingly turn to LLMs for medical advice, which model they use and what words they choose could lead to drastically different informal triage guidance.^17^

Of particular note were differences in responses to “weak” versus “numb.” Although the CDC highlights both as stroke symptoms, only Llama treated them equally.^5^ Both Claude and GPT-4o more aggressively sought care for “weak” than “numb,” suggesting a mismatch between public health messaging, patient language, and model behavior. Future work should focus on understanding how individuals experiencing a CVA describe their symptoms. Interestingly, no model ever chose to drive to the ER, despite this being a common real-world choice, a deviation with important implications for using LLMs to model human behavior.^6,17^

This study had several limitations. Commercially available LLMs are constantly evolving, so these comparisons may represent a moment-in-time snapshot rather than persistent model-to-model differences.^18^ LLM behavior is also sensitive to prompt architecture, and alternative prompts could yield different patterns.^19,20^ Future work should expand to different prompt structures and evaluate native human-AI interactions. Additionally, this study examined LLM responses to hypothetical scenarios, not actual human behavior. Care-seeking decisions are multifactorial, and how individuals incorporate LLM advice into those decisions is beyond this study’s scope.^6,9,21^

Ultimately, as people turn to LLMs for informal medical advice, understanding how these models recommend accessing and utilizing medical care becomes critical. In this study, LLMs frequently provided poor and inconsistent triage advice for stroke, findings that are concerning both for individuals who may rely on them when facing a potential stroke, and for broader public health efforts to improve stroke recognition and early 911 activation.

## Data Availability

All data produced in the present study are available upon reasonable request to the authors

## Notes

**Disclosure statement:** The authors have no relevant conflicts of interest to disclose.

### Competing Interest Statement

The authors have declared no competing interest.

